# Automated Model Discovery Based on COVID-19 Epidemiologic Data

**DOI:** 10.64898/2026.02.22.26346850

**Authors:** Morteza Babazadeh Shareh, Florian Kleiner, Michael Böhme, Corinna Hägele, Petra Dickmann, Rainer Heintzmann

**Affiliations:** Leibniz Institute of Photonic Technology, Jena, _Thuringia_, Germany; Institute of Inorganic and Analytical Chemistry, Friedrich Schiller University Jena, _Thuringia_, Germany; Institute of Physical Chemistry and Abbe Center of Photonics, Friedrich Schiller University Jena, Jena, _Thuringia_, Germany; Department of Anaesthesiology and Intensive Care, Jena University Hospital, Jena, Germany; Leibniz Centre for Photonics in Infection Research (LPI), Jena, Germany

**Keywords:** COVID-19 Pandemic, Epidemiological Modelling, Sparse Identification of Nonlinear Dynamical Systems (SINDy), Automated Model Discovery, Data-Driven Modelling, Universal Differential Equations (UDE), Infectious Disease Dynamics

## Abstract

The COVID-19 pandemic has presented severe challenges in understanding and predicting the spread of infectious diseases, necessitating innovative approaches beyond traditional epidemiological models. This study introduces an advanced method for automated model discovery using the **S**parse **I**dentification of **N**onlinear **Dy**namics (SINDy) algorithm, leveraging a dataset from the COVID-19 outbreak in Thuringia, Germany, encompassing over 400,000 patient records and vaccination data. By analysing this dataset, we develop a flexible, data-driven model that captures many aspects of the complex dynamics of the pandemic’s spread. Our approach incorporates external factors and interventions into the mathematical framework, leading to more accurate modelling of the pandemic’s behaviour. The fixed coefficient values of the differential equation as globally determined by the SINDy were not found to be accurate for locally modelling the measured data. We therefore refined our technique based on the differential equations as found by SINDy, by investigating three modifications that account for recent local data. In a first approach, we re-optimized the coefficient values using seven days of past data, without changing the globally determined differential equation. In a second approach, we allowed a temporal dependence of the coefficient values fitted using all previous data in combination with regularization. As a last method, we kept the coefficients fixed to the original values but augmented the differential equation with a small neural network, locally optimized to the data of the past week. Our findings reveal the critical role of vaccination and public health measures in the pandemic’s trajectory. The proposed model offers a robust tool for policymakers and health professionals to mitigate future outbreaks, providing insights into the efficacy of intervention strategies and vaccination campaigns. This study advances the understanding of COVID-19 dynamics and lays the groundwork for future research in epidemic modelling, emphasising the importance of adaptive, data-informed approaches in public health planning.

## Introduction

The global outbreak of COVID-19 has underscored the critical importance of timely and accurate modelling in understanding and combating infectious diseases. Traditional mathematical models, such as the Susceptible-Infected-Recovered (SIR) model, have provided foundational insights into disease dynamics [1, 2, 3]. However, the complexity and unprecedented nature of the COVID-19 pandemic have revealed limitations in these classic approaches, particularly in their adaptability to rapidly changing data and their capacity to capture the complex interactions within and between populations [4]. In response to these challenges, we introduce the application of the Sparse Identification of Nonlinear Dynamical Systems (SINDy) algorithm [5] performing automated model discovery based on COVID-19 data. This algorithm starts with a matrix of the potential terms (called basis) contributing to the desired differential equation model. By enforcing sparsity, it then determines a minimal set of required terms along with their coefficients. In this way, SINDy can extract the best differential equation approximating the system consistent with the measured data. Leveraging the power of this algorithm, we aim to transcend the constraints of conventional models, offering a more flexible and data-driven pathway to deciphering the complex dynamics of the disease spread. In the rest of the section, we review previous approaches to COVID-19 modelling.

The limitations of traditional compartmental models have prompted researchers to explore enhancements by incorporating artificial intelligence elements. For example, in one COVID-19 modelling project, Alqahtani explores an innovative extension of the traditional SIR compartmental model by integrating fractional derivatives [6]. This approach offers a nuanced understanding of the epidemic’s dynamics by incorporating memory effects, providing a more detailed analysis of infection spread over time. Alqahtani’s work demonstrates the potential of modifying classic models to enhance their descriptive power, stability, and numerical analysis capabilities. This work underscores the importance of evolving mathematical frameworks to deal with the complex characteristics of the COVID-19 pandemic, setting a good example for using advanced tools, such as the SINDy algorithm, for automated model discovery. In another study, Vega et al. made a significant leap towards integrating machine learning algorithms with the SIR epidemiological model. Their development of the SIMLR framework incorporates machine learning to refine COVID-19 forecasts, offering a novel method that enhances prediction accuracy by learning from the data in real-time [7]. This hybrid approach illustrates the potential of machine learning to augment traditional models and sets a crucial groundwork for further explorations into adaptive, data-driven modelling techniques. Similarly, Kong et al. introduce a hybrid modelling strategy that combines the predictive strengths of epidemic differential equations with the adaptive learning capabilities of recurrent neural networks (RNNs). This method significantly advances the accuracy of COVID-19 prevalence forecasts and shows the potential of hybrid models in navigating the complexities of pandemic dynamics [8]. By leveraging the structured insights of epidemic models with the flexible, data-driven nature of recurrent neural networks (RNNs), their work paves the way for more resilient forecasting tools.

Recent research aims to shed light on the importance of the interventions and external factors in the dynamics of the pandemic. In one study, Dehning et al. contributed significantly to the understanding of the temporal dynamics of COVID-19 through the lens of change-point-analysis. By accurately analysing the effects of public health interventions on the pandemic’s trajectory, this research offers insights into how timely and targeted measures can alter the course of the disease’s spread. The study provides a quantitative framework for evaluating intervention strategies and highlights the critical need for adaptable and data-informed models in public health planning [9]. Another study has comprehensively explored the complicated role of epidemiological models in analysing the complex dynamics of the COVID-19 pandemic [10]. This research uses modelling to monitor and predict the outbreak’s trajectory, evaluating the efficacy of public health interventions, and informing policy decision makers. By synthesising data across various scales and contexts, the study highlights the essential value of epidemiological models in offering insights critical to shaping a coordinated and effective response to the pandemic. A pivotal study [11] explores the global impact of vaccine distribution strategies using an advanced version of the SIR model. By analysing data from 152 countries in 2021, the potential benefits of equitable vaccine sharing in reducing the global burden of COVID-19 were described. Their findings underscore the importance of prioritising need over wealth in vaccine distribution to mitigate the spread of the virus and advocate for equitable solutions in the pandemic response. The studies above illustrate the importance of considering external factors in pandemic modelling. The ability of the SINDy algorithm to accept control signals as external factors helps us incorporate such external factors [5].

Given the limitations of the current mathematical pandemic models, some researchers are trying to develop methods based on purely data-driven approaches. For example, in [12], the authors present a novel approach to forecast virus outbreaks by leveraging social media data and neural ordinary differential equations (NODEs). By integrating real-time information from social media platforms into their modelling framework, the researchers demonstrate the potential for improved epidemic forecasting accuracy. Their use of NODEs allows for dynamic modelling of complex, nonlinear interactions, enabling more precise predictions of outbreak trajectories. This innovative methodology highlights the valuable insights that can be gathered from unconventional data sources and underscores the importance of flexible modelling approaches in addressing dynamic public health challenges. However, their method relies heavily on recent data (two months) and does not yield a globally valid model of the underlying epidemic dynamics. Gao et al. introduce an evidence-driven spatiotemporal prediction model leveraging Ising dynamics [13] for COVID-19 forecasting. By integrating diverse data sources and employing Ising dynamics, a statistical physics approach, their model provides insightful predictions of COVID-19 hospitalisations at both spatial and temporal scales. This methodology accounts for complex interactions between various epidemiological factors and geographical regions, offering a nuanced understanding of disease dynamics. The study’s emphasis on evidence-driven predictions highlights the importance of data-driven approaches for effective public health interventions. Sandie et al. present an analysis of the observed versus estimated trends in COVID-19 case numbers in Cameroon [14]. They use a statistical technique, Multilevel Regression with Poststratification (MRP), and then apply Seasonal Autoregressive Integrated Moving Average (SARIMA) analysis to provide valuable insights into the pandemic dynamics. By comparing actual case data with model predictions, they evaluate the forecasting accuracy of existing modelling frameworks. This research sheds light on the challenges and limitations inherent in pandemic modelling, particularly in resource-constrained settings. From these studies we learn that refined modelling techniques and incorporating local data to improve the accuracy of disease forecasts are essential.

All of these studies show that the current mathematical models (SIR family) cannot handle the complexity of the COVID-19 pandemic, calling for improvements. Despite making valuable contributions, these methods still suffer from the limitations of the traditional hand-crafted mathematical models requiring postprocessing. In contrast, we aim to automatically extract powerful mathematical models capable of capturing the complex features of the pandemics directly from the data using SINDy. We hope that this creates mathematical models that inherently represent the pandemic system, eliminating the need for auxiliary methods to enhance model performance. We apply a data-driven approach to a local dataset aiming to extract the mathematical model automatically. Using a data-driven approach avoids making prior assumptions about the role of different variables in the system before constructing the model. We instead let the model discovery algorithm analyse the (Thuringian) dataset to construct a suitable pandemic model.

In this work, we use some pre-processing methods to reduce the noise and make the data more suitable for modelling. Moreover, we apply optimisation methods to make the final model more accurate and capable of capturing temporal external factors. The rest of the paper is organised as follows: First, we explain the method, then some results from the proposed method, and finally, in the conclusion section, we discuss our work.

## Methods

Figure 1 provides an overview of the methodological framework employed in this study, which begins with a pre-processing stage, followed by mathematical modelling and model optimisation. In the following, we first describe the data, then present a high-level description of the method, and finally detail each step of the pipeline.

**Figure 1.**
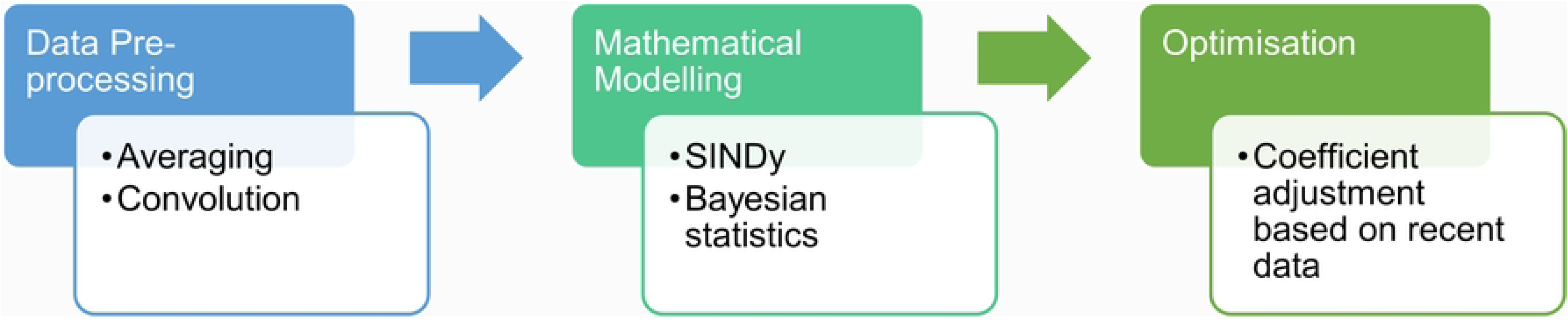
Overview of the proposed model. See text for more details on the various steps.

### Data

The primary time-series dataset was derived from COVID-19 patient records gathered by the health authorities of the Free State of Thuringia and curated by the Pandemics Control Group, starting on the 3^rd^ of March 2020 (the first SARS-CoV-2 case in Thuringia, Germany) and ending on the 7^th^ of February 2022. Thuringia is a federal state in central Germany with about 2.1 million inhabitants and a relatively high average age of around 47 years, according to the Thuringian State Office for Statistics (TLS, 2022). The raw dataset comprised roughly 400,000 anonymous records from patients in various districts. Every record contained detailed information such as infection date, age, gender, vaccination, and hospitalisation status. This information was processed to build a comprehensive time-series dataset of infection, hospitalisation and admission to the intensive care unit (ICU) stratified by age group. This primary dataset covers more than two years of the pandemic records. However, the quality of these data must be considered with caution. Actual infection numbers were likely underestimated, particularly during peak infection periods when registration offices were overwhelmed, and in later stages of the pandemic when many individuals chose not to report their infections. The effect of underreporting is especially noticeable around Christmas and New Year, where a sharp decline in reported cases can be observed. Another set of information we used in the model is the overall vaccination records in Thuringia. This data starts on the 27^th^ of December 2020 (first jab in Thuringia) and ends on the 30^th^ of June 2022. The raw vaccination data, gathered by the health authorities of the Free State of Thuringia and curated by the Pandemics Control Group, recorded all vaccination centre activities in the state. Each centre recorded on a daily basis how many people, at which age, got vaccinated and whether it was their first, second, or third jab. After processing these data, we built a comprehensive time-series dataset describing all vaccinations in the state.

### Method description

We aim to predict the number of infections, hospitalisations, and ICU cases for a specific time range. The main approach of the proposed method is to develop a model that captures the dynamic patterns of the pandemic, including its peaks, declines, and fluctuations over time. As Figure 1 shows, the proposed method has three steps: we first pre-process the data to reduce the noise in the dataset and extract two new features (Infectiveness and Antibody) using the measured information. The reported case numbers showed a clear dependence on the day of the week of reporting, which we corrected using a sliding-window weekly average. This single weekly averaging step also mitigated delays in reporting during holiday periods, particularly around Christmas and Easter. In the pre-processing, we also convolved the data to extract two new features (Infectiveness and Antibody). The second step was to automatically discover a mathematical model representing the dynamics of COVID-19 infections in Thuringia. This mathematical model has two parts. The first part consists of a number of ordinary differential equations extracted from the time-series dataset. These ODEs modelled only the number of infections. We used the SINDy algorithm to discover these equations. The second part of the mathematical model aims to predict the number of hospitalisation and ICU cases. Since we observed a linear correlation between hospitalisation (also ICU) and infection, Bayesian statistics were used to implement a probabilistic regression on the data. The third and final step of our method is to optimise the coefficients in the differential equations based on more recent time stretches of the data (one week), to allow better predictions. The extracted ODEs using SINDy provided an overall model of the pandemic’s non-linear behaviour. Yet, when we want to make a projection from a particular starting date, we expect that external temporary confounding factors (like a period of public health intervention) may have significant influence on the dynamics. Therefore, when projecting from a certain date into the future, we first optimised the coefficients of the ODEs based on one week of data points before that date. In this way, such recent influences on the pandemic were accounted for. In the following section, every step will be discussed in detail. Table 1 lists the symbols used in the equations throughout this section.

**Table 1.**
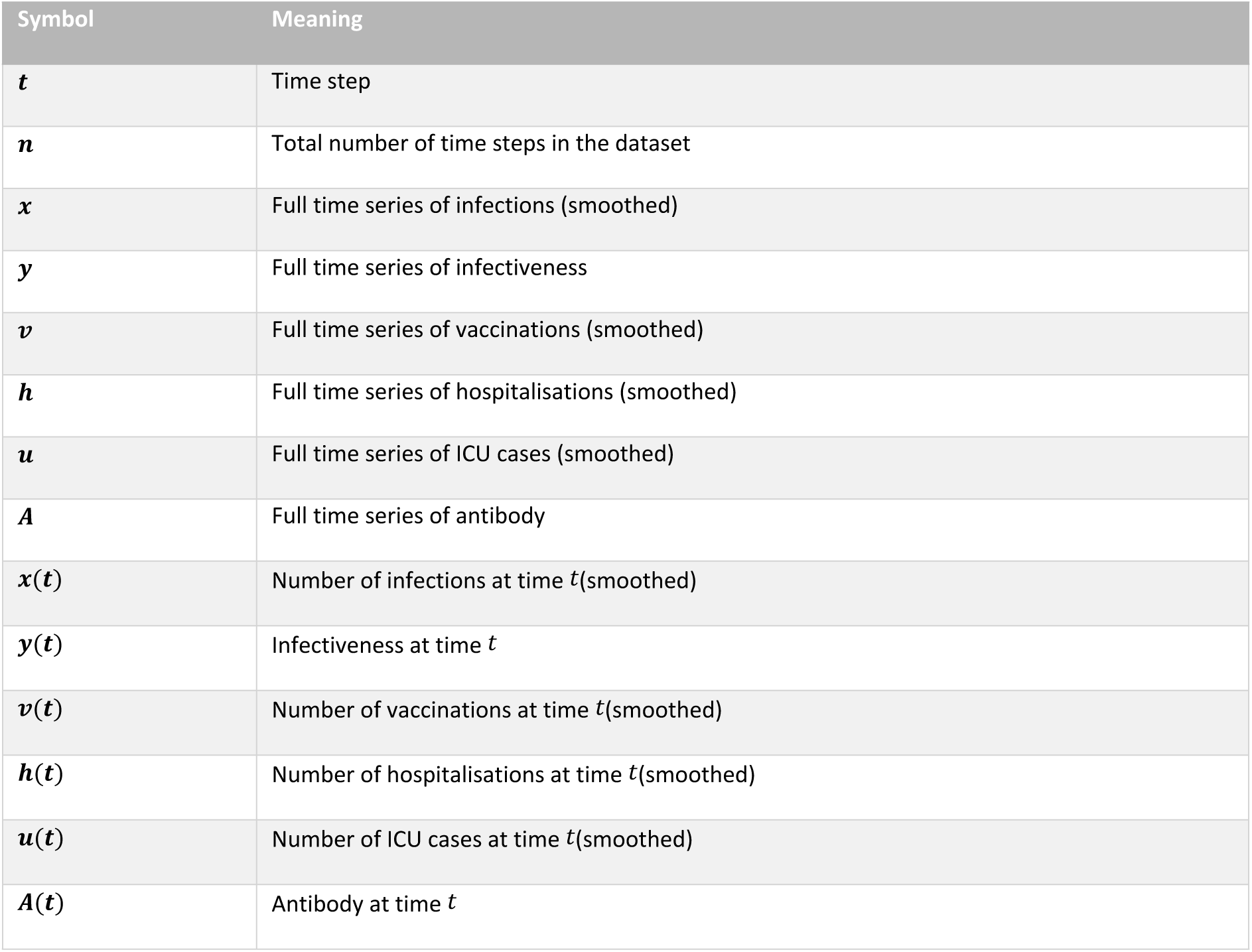
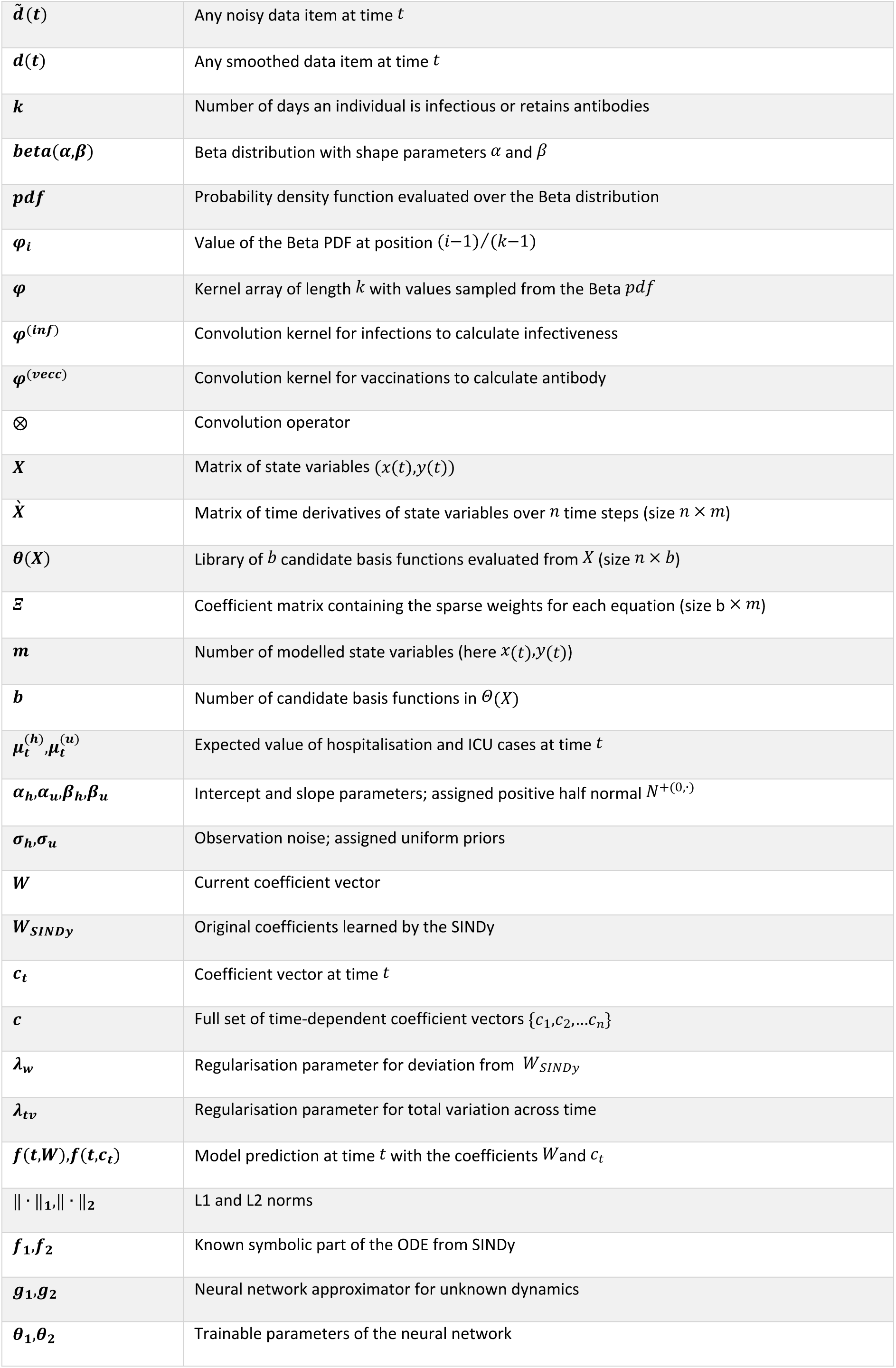
Definitions of variables, parameters, and functions used in the Methods section.

### Step 1: Data pre-processing

The dataset must first be pre-processed to become suitable for the SINDy algorithm. Two different issues in the dataset need to be addressed before the next step. One of the critical problems in the raw dataset is biases in reporting infections, hospitalisations, ICU cases and vaccinations. Since many healthcare offices were closed on weekends, the weekend cases were often reported in the first days of the following week. This reporting bias caused strong fluctuations in the data. To overcome this problem, we smoothed the data. According to the nature of the reporting bias, which varied with the day of the week, a weekly averaging box-filter of uniform weight (Eqn. 1) was applied to the raw data, where *d~(i)* is any noisy data (infections, hospitalisations, ICU cases and vaccinations) reported at time *t*. The output *d(t)* is the smoothed version of this data. Here, *n* is the total number of data points in the time series data. The number six in the equation shows that it is a weekly averaging. Throughout this paper all variables (e.g. *x(t), u(t)*) refer to smoothed data series unless explicitly marked with a tilde (e.g. *x~(t)*) to indicate raw data.

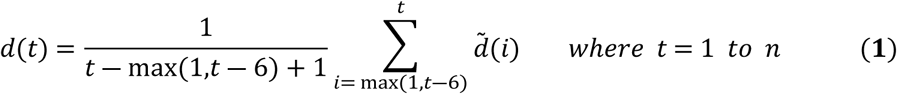

Since SINDy discovers only ordinary differential equations (ODEs) without hidden states, it cannot directly represent delays, such as the time between infection and the onset of infectiveness. To account for such delays, we incorporated them during the pre-processing step by convolving the measured time series (infections and vaccinations) with appropriate kernel functions. For each kernel, we chose a Beta distribution, which is defined by two shape parameters, *α* and *β*, and allows us to generate flexible bell-shaped curves [15]. Equation (2) defines a *β* -distribution based kernel function denoted as *ϕ*.

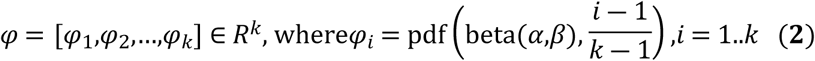

The vector *ϕ* consists of *k* values sampled from the Beta distribution’s probability density function (PDF), where *k* is the number of days during which an infected person is considered infectious, or a vaccinated person retains antibodies. The kernel *ϕ* captures how infectiveness or antibody levels evolve over time after the respective event. Each point *ϕ_i_*corresponds to the value of the Beta PDF evaluated at the normalized position *(i-1)/(k-1)*, ensuring the kernel spans the full interval *[0,1]* and forms a smooth curve suitable for convolution.

Equations (3) and (4) show how the convolution is carried out on the data. *Φ^(inf)^* is the kernel used to convolve the infections and *ϕ^(vacc)^* is for the vaccinations. *x* and *v* are the full smoothed time series of the infections and vaccinations data respectively, and the outputs *y* and *A* we call “infectiveness” and “antibody”.

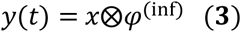

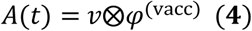

### Step 2: Mathematical Modelling

The output of the previous step is a smoothed version of the dataset, enriched with additional informative features (the infectiveness and antibody). In this step, we construct a mathematical model based on the pre-processed time-series data. Our goal is to predict infections, hospitalisations, and ICU cases. Modelling all three variables simultaneously was found to be too complex and potentially underdetermined, so we adopted a hierarchical approach. We first use the SINDy algorithm to identify the underlying differential equations governing the infection dynamics. Based on the observed linear correlation between infections and subsequent hospitalisations (including ICU admissions), we then use the predicted infections to estimate the other variables.

We begin by presenting the SINDy-based modelling of infections, followed by the prediction of hospitalisations and ICU cases. Equation (5) presents the matrix sparsification framework used by SINDy to identify a compact system of differential equations that best captures the infection dynamics.

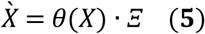

Equation (5) expresses the core structure of the SINDy algorithm as a sparse regression problem. The matrix *Ẋ* ∈ ℝ*ⁿˣᵐ* contains the numerically computed time-derivatives of the state variables (namely infections *x(t)* and infectiveness *y(t)*) evaluated over *n* time steps. The matrix *θ(X)* ∈ ℝ*ⁿˣᵇ* is the library of the *b* candidate basis functions constructed from the state variables and the external input *A(t)*, which represents the antibody level. Although *A(t)* is not part of the state dynamics being directly modelled, it is included as a control signal influencing the system. The matrix *Ξ* ∈ ℝ*ᵇˣᵐ* contains the sparse coefficients that determine the contribution of each basis function to the differential equations governing *x(t)* and *y(t)*. Here, *n* is the number of time points, *m* is the number of modelled variables, and *b* is the number of basis functions in the library.

To estimate *Ξ*, the SINDy algorithm solves a sparse regression problem using the Sequentially Thresholded Least Squares (STLSQ) method. This method includes a threshold parameter *λ* which controls the sparsity of the solution. A lower *λ* yields a sparser model with fewer active terms but may reduce accuracy, whereas a higher *λ* results in a more complex model that better fits the data. We solved the problem by using different values of *λ* and selected the Pareto-optimal solution as the final model.

After identifying a suitable model for infections via SINDy, the second part of the modelling process focuses on predicting the number of hospitalisations and ICU cases. Because a strong correlation between hospitalisations (and ICU cases) and infections is expected, we use a linear model based on probabilistic regression. Bayesian statistics are employed to account for uncertainty. Equation (6) shows how hospitalisations and ICU cases are estimated from the infection data:

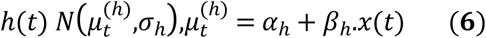

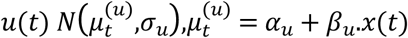

Equation (6) defines a probabilistic linear regression model used to predict hospitalisations, *h(t)*, and ICU cases, *u(t)*, based on the number of infections, *x(t)*, at time *t* and *N(μ, σ)* denotes the normal distribution with expectancy *μ* and standard deviation *σ*. Each of the two observed outcomes is assumed to follow a normal distribution, centred at a linear mean function: *μ⁽ʰ⁾ = α + β · x(t)* for hospitalisation, and *μ⁽ᵘ⁾ = αᵤ + βᵤ · x(t)* for ICU cases. The parameters *α* and *β* represent the intercept and slope of each linear relationship and are assigned positive half-normal priors *N⁺⁽⁰, ·⁾* to enforce nonnegativity. The noise parameters *σ* and *σ_u_* capturing the uncertainty in observations, are drawn from uniform distributions. This Bayesian regression framework enables uncertainty-aware prediction of hospitalisation and ICU trends based on infection counts. This model is applied across all time points *t = 1, 2, …, n*, allowing the regression parameters *α*, *β* and *σ* to be inferred from the full infections and hospitalisations (or ICU cases) time series.

### Step 3: optimization

The system of differential equations obtained via SINDy provides a global model of the pandemic dynamics, incorporating the antibody level as an external control input. However, the model coefficients may also be influenced by unobserved or time-varying external factors, such as policy changes or shifts in public behaviour. As a result, while the SINDy model captures the overall trends of the system, discrepancies between its predictions and the observed data can still occur.

To address these deviations, we allow the coefficients of the ODEs to adapt over time in response to such external influences, while keeping the structural form of the equations fixed. For this purpose, we applied three different optimisation strategies, each designed to fine-tune the model parameters and improve predictive performance under dynamically changing conditions. These strategies are referred to as:

1- Local coefficient adjustment,
2- Time-dependent coefficient adjustment, and
3- Neural-augmented ODE adjustment.

We refer to the original, unoptimised coefficients obtained directly through SINDy as the global coefficients. The details of each optimisation strategy are presented in the following paragraphs.

Equation (7) defines the local coefficient adjustment approach to optimise the coefficients *W* of the SINDy-based model for improved short-term prediction. The objective function minimises the squared error between the model output *f(t,W)* and the observed infection data *x(t)* over the most recent *K* time steps, ending at the prediction date *D*. A regularisation term, weighted by *λ_w_*, penalises deviations from the original SINDy coefficients *W_SINDy_*, ensuring that the locally optimised model remains close to the globally learned dynamics.

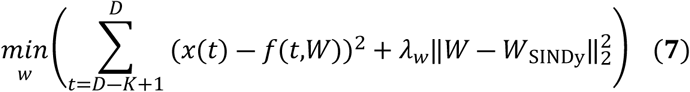

Equation (8) shows the time-dependent coefficient adjustment approach, which allows the models coefficients *c_t_* to change over time in response to external influences. The objective function minimises the squared error between the model output *f(t, c_t_)* and the observed infections data *x(t)* over all time steps up to the prediction date *D*. To ensure temporal smoothness and prevent overfitting, a total variation regularisation term is added. This term penalises abrupt changes between consecutive coefficient vectors *c_t_* and *c_t-1_*, scaled by the regularisation parameter *λ_tv_*. In this approach, to make a projection from a specific date *t*, the corresponding coefficient vector *c_t_* at that time is used in the model.

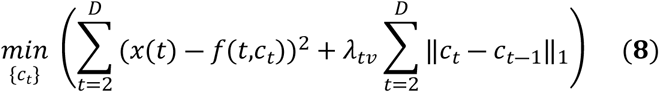

Equation (9) defines the neural-augmented ODE adjustment approach, in which the original differential equations obtained by SINDy are extended with neural networks to capture additional time-dependent influences not explicitly modelled. The functions *f_1_* and *f_2_* represent the deterministic dynamics learned from the SINDy model, while the neural networks *g_1_*and *g_2_*, parameterised by *θ_1_* and *θ_2_*, are trained to approximate unknown external factors. All components are evaluated using the time-dependent variables *x(t)*, *y(t)* and *A(t)* where *x(t)* and *y(t)* are infections and infectiveness respectively and *A(t)* is the antibody level as a control input. The neural terms allow the model to adapt to dynamic effects without changing the original ODE. This approach is implemented using the Universal Differential Equations (UDE) framework, which enables the hybrid integration of mechanistic and data-driven components [16].

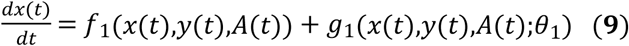

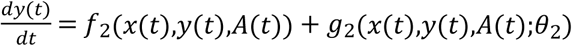

This study used only aggregated and fully anonymized epidemiological surveillance data provided by the Thuringia state health authorities. The dataset contained no individual-level records or variables that could allow identification of natural persons. The aggregated data used for this analysis were first accessed by the corresponding author on 01 October 2021. No human participants were recruited, and no personal or medical-record data were accessed. Because the study did not involve human subjects or identifiable human participant information, ethics committee approval and informed consent were not required.

In the next section, we will look at some results using the proposed method.

## Results

### Dataset Illustration

The proposed model has been designed and developed based on Thuringia’s COVID-19 data, which covers 707 days of the pandemic. The raw reported data was processed and transformed into a time series dataset using the DataFrames.jl package [17] in the Julia programming language. This time-series dataset provides quite a detailed account of the progress of the epidemiological spread of the pandemic in the German state of Thuringia. For example, the smoothed version of infections for different age groups (left panel) and hospitalisations (and ICU cases) for 60-79 age-group (right panel) are summarized in Figure 2. All reported results are based on this Thuringia dataset as the input.

**Figure 2.**
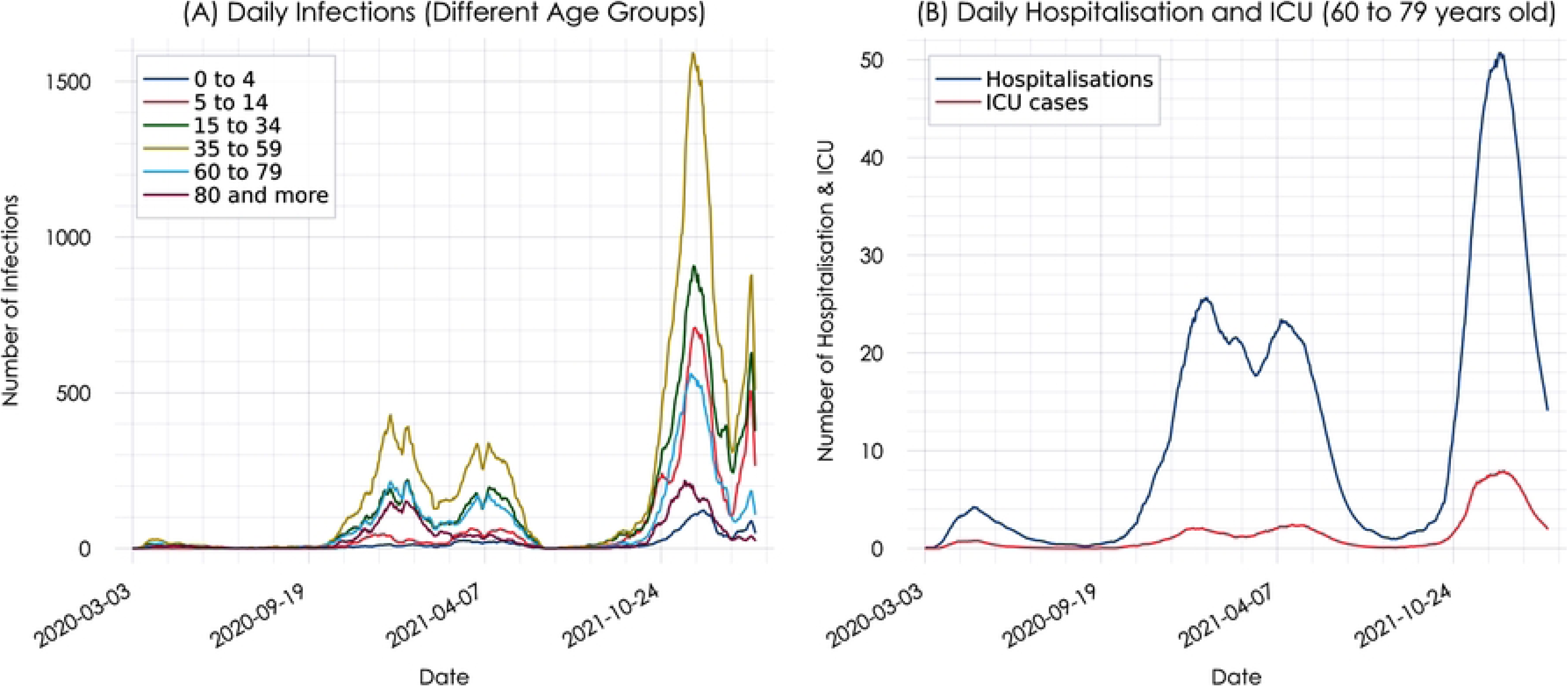
Some details of the Thuringia time-series dataset. The left panel (A) shows the measured infections per day for various age groups. The right panel (B) displays the hospitalisations and ICU cases (ages 35-59) attributed to COVID-19.

The plots shown in Figure 2 are based on weekly-averaged numbers. The same averaging process has been applied to the vaccinations data (Figure 3). The difference between the two plots shows the effect of weekly averaging (Fig. 3, right panel) on noisy reported data (Fig. 3, left panel).

**Figure 3.**
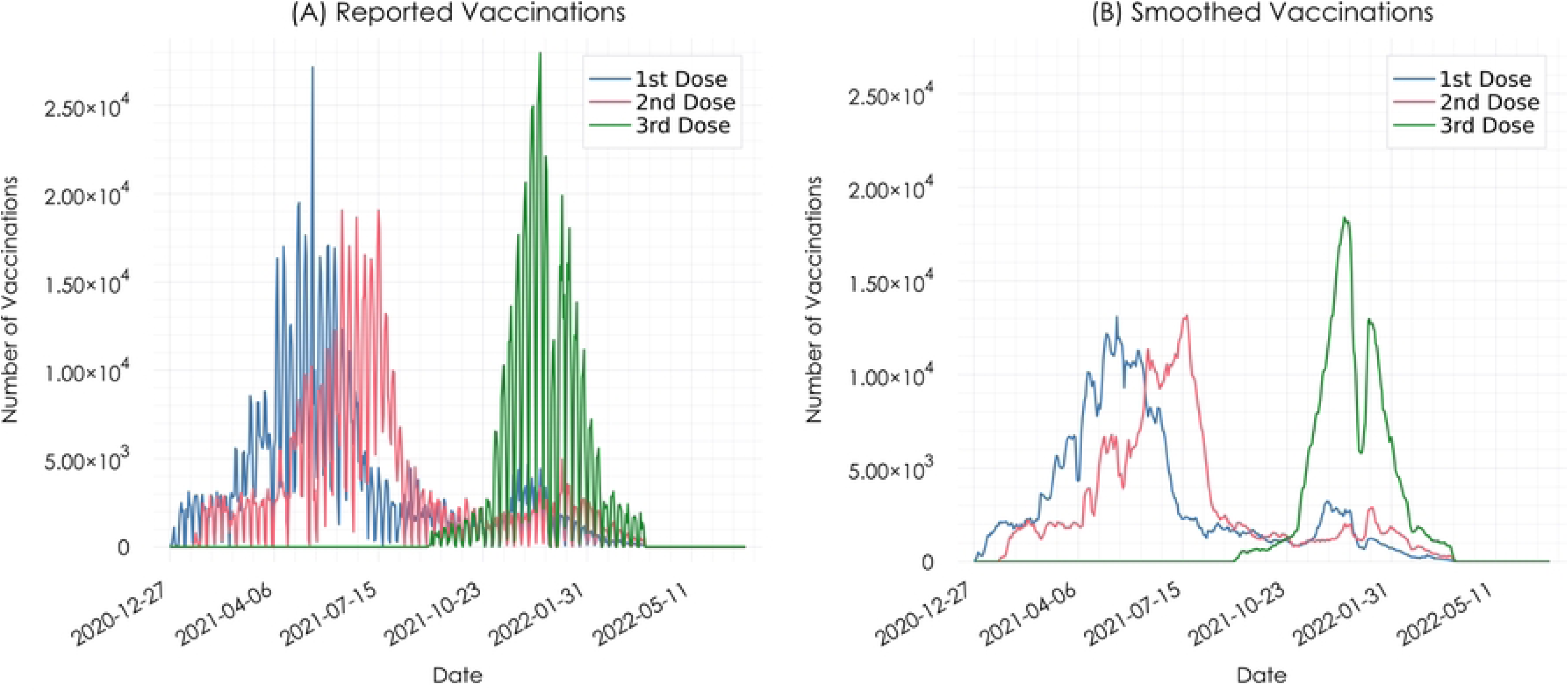
Vaccinations before and after averaging. On the left panel (A) the daily statistics is shown. On the right (B), only the weekly averaging is displayed.

### Antibody and infectiveness

Two kernel functions have produced two new features, antibody (known as *A*) and infectiveness (known as *y*) through convolution. Equations (3) and (4) in the previous section show how we calculate them. For the antibody kernel, different articles claim various antibody persistence durations for COVID-19 vaccines [18, 19]. However, through validation, we noticed that a 35-day period with the highest antibody after two weeks gives the highest quality antibody values for the model. The infectiveness kernel was designed based on the duration of the COVID-19 disease. According to many articles, COVID-19 lasts for three weeks, and at the end of the first week, the level of ability to infect others culminates [20, 21]. Therefore, we used this kernel to extract infectiveness. Both kernels were produced using the Julia programming language’s Distributions.jl package [22]. Figure 4 shows the kernel functions and the results after convolution. All numbers are normalised to the [0,1] range. A closer look at the green lines on the Antibody1 plot (first dose) shows that despite having the same number of vaccinations on the 110^th^ and the 170^th^ day (see dotted green vertical lines), the level of antibody is much higher on the 170^th^ day due to the different previous vaccination history. The same delay rule applies to the 620^th^ and 660^th^ day on the infectiveness plot. These convolved signals will be further used in the mathematical models.

**Fig 4.**
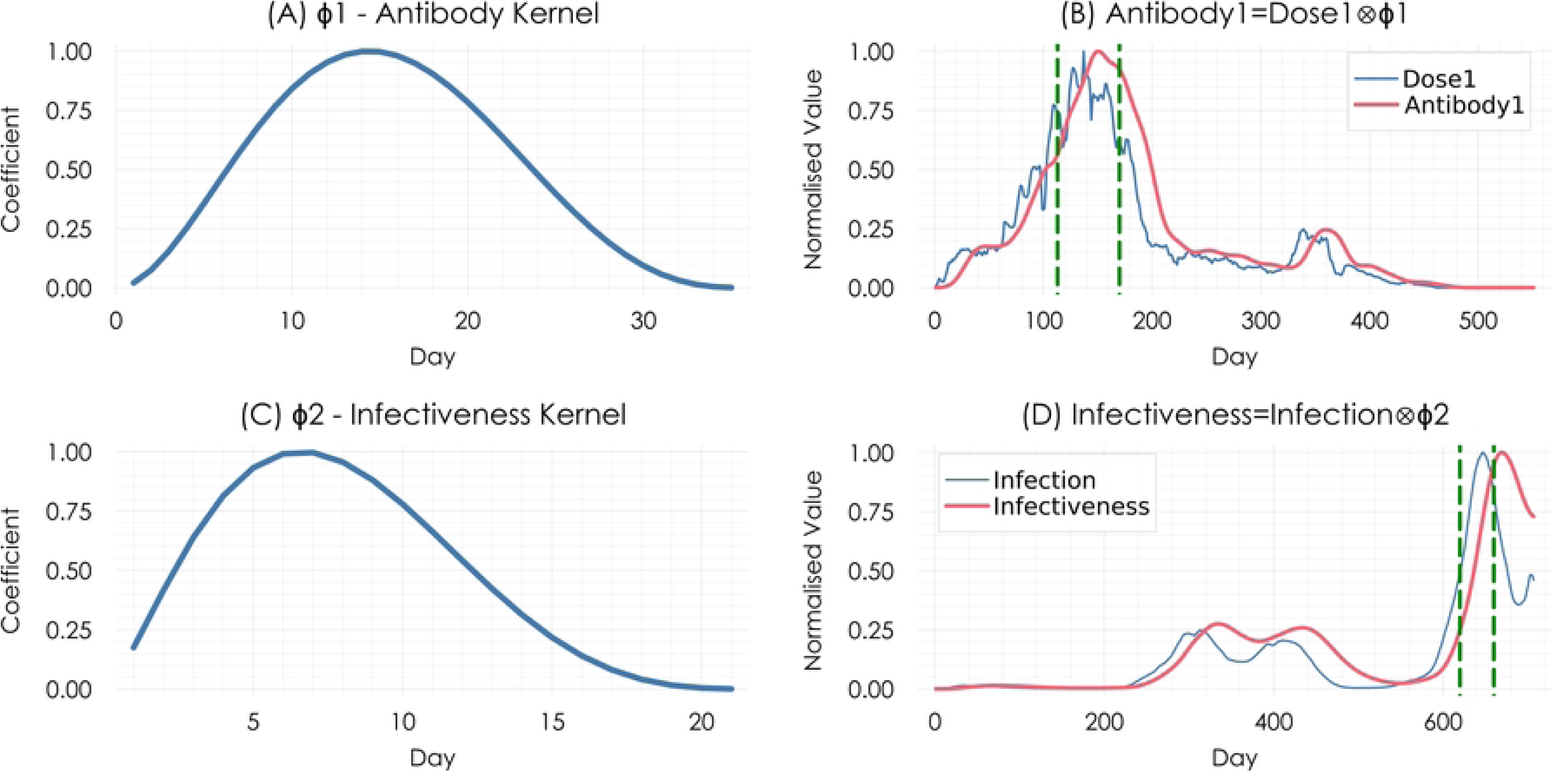
Antibody and infectiveness kernels. The left plots (A and C) show the kernel functions, while the right plots (B and D) illustrate the effect of convolution on vaccinations and infections using these kernels.

### Mathematical model for infections

The next step after pre-processing is developing a mathematical model for the data. Sparse Identification of Non-linear Dynamic (SINDy) has been used to create a model of the infection dynamics. It is an automated model discovery approach. The parameters used in this research are specified in Table 2.

**Table 2.** The parameters of the SINDy.

| Parameter | Value |
| --- | --- |
| Type of the Problem | Continuous |
| Variables | infections, infectiveness ($x, y$) |
| Control Signal | antibody ($A$) |
| $\lambda$ | $[10^{-8}, 10^{-1}]$ |
| Basis | Polynomial |

As it is evident from Table 2, the algorithm’s input comprises two variables, infections and infectiveness (called *x, y*) as well as antibody (called *A*) as control signal. Antibody and infectiveness are the features extracted from the vaccinations and infections, as explained in the previous section. The range of *λ* that we use in the model discovery process is *[10^-8^, 10^-1^]*. *λ* is the regularisation parameter in the SINDy method, controlling the balance between fitting the observed data and sparsity. A higher value of *λ* encourages sparser models, meaning fewer terms are selected in the identified equations. Conversely, a lower value of *λ* allows for more terms to be included in the model, potentially capturing more complexity in the system dynamics. We used a range of *λ* to find a Pareto front on the solution space. The DataDrivenDiffEq.jl package [23] was used to discover a mathematical model. The SINDy algorithm is part of this package. According to the designed model, we can exclude any part of the data, make a model with the rest, and then run a prediction for the excluded part. For the prediction, we solve the differential equations numerically using the DifferentialEquations.jl package [24] in the Julia language.

Employing stochastic differential equations (SDEs) instead of ordinary differential equations (ODEs) allows for a more comprehensive representation of chaotic systems, such as the COVID-19 pandemic, by accounting for inherent uncertainties. While ODEs yield a single deterministic trajectory, SDEs generate a distribution of possible outcomes through iterative simulations. The resulting spread reflects the system’s uncertainty, with the central tendency indicating the most probable evolution. Since the SINDy algorithm produces a deterministic ODE system, we augmented the discovered model with a diffusion term to formulate a corresponding SDE. The magnitude of this diffusion term was determined empirically through iterative testing, selecting a value that balanced model stability and data fit. All projection results in this section were computed using the stochastic version of the model to account for uncertainty

Figure 5 shows three examples of daily and weekly predictions after solving the SDE 5000 times. The plots on the top show a one-month prediction, the middle ones predict 45 days and the bottom plots make predictions for two months. A different range of dates was used in these examples. In the daily prediction plots, the most likely prediction is the blue line, but the ribbons specify other ranges of possibilities. Since the model was created using smoothed data, having a weekly prediction is necessary to make it helpful. The boxplots on the right side show the equivalent weekly predictions.

**Fig 5.**
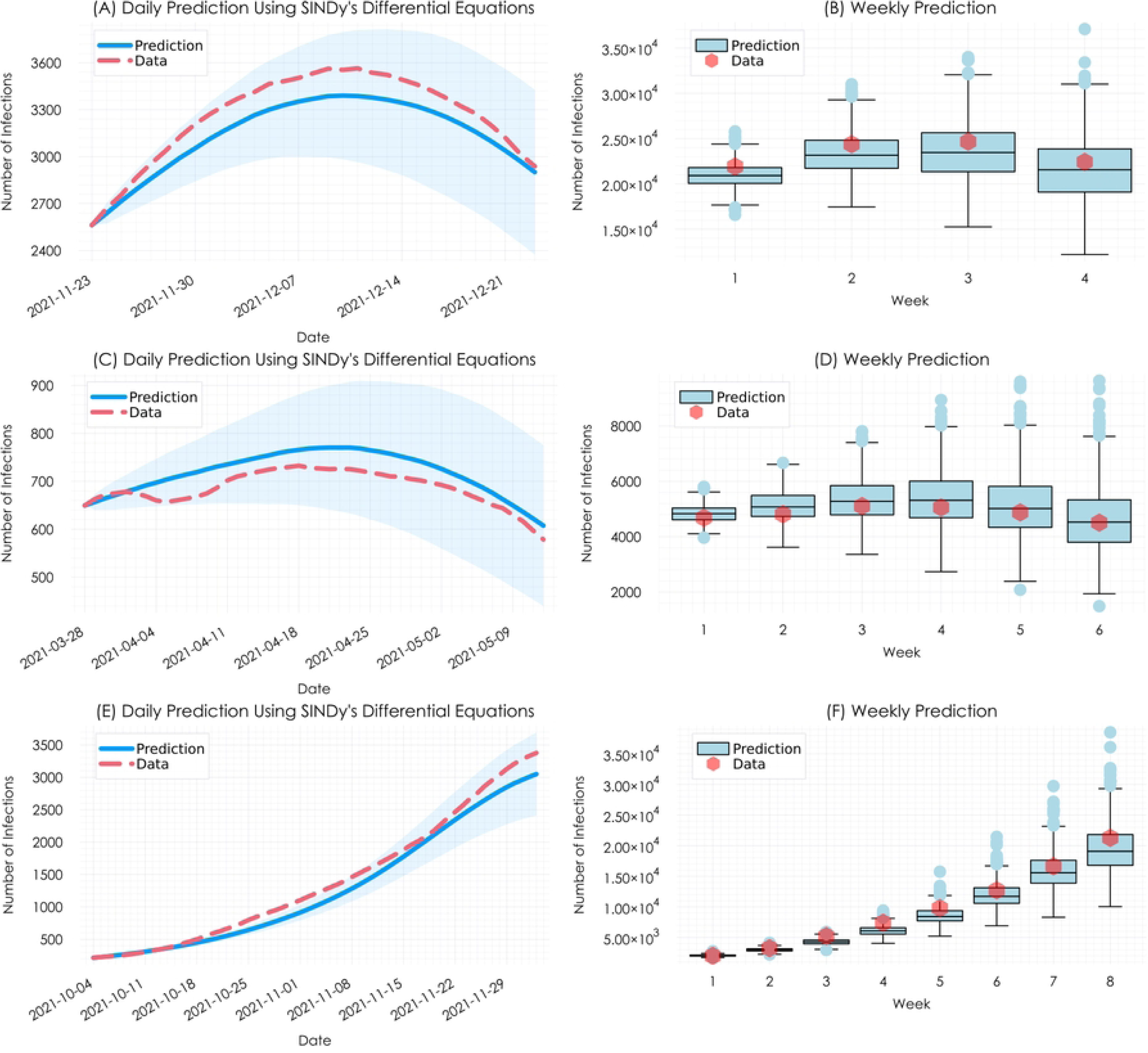
Daily and weekly predictions. The plots show predictions for one month (A and B), 45 days (C and D), and two months (E and F) across different time frames.

### Hospitalisations and ICU cases

Since there is a correlation between infections and hospitalisations (and ICU cases), linear regression was used to predict the number of hospitalisations (and ICU cases). As shown in the previous section, it is possible to predict the number of infections through the discovered mathematical model using SINDy. Now, we can use these infection numbers to forecast hospitalisations and ICU cases. To account for uncertainty, Bayesian statistics were used. To ensure the model captures the most recent trends in the correlation between infections and hospitalisations (and ICU cases), only the 60 data points prior to the prediction day were used to train the Bayesian model. A probabilistic regression was applied to predict the number of hospitalisations (and ICU cases). The posterior distribution (for hospitalisations) after 1,000 iterations with the No-U-Turn Sampler or NUTS is shown in Figure 6. It is carried out according to Equation (6). The Turing.jl package [25] was used for probabilistic programming. The first 500 iterations were used to stabilise the stochastic model. *α_h_*, *β_h_* and *σ_h_*in the plots represent intercept, slope, and observation noise of the hospitalisations, respectively.

**Figure 6.**
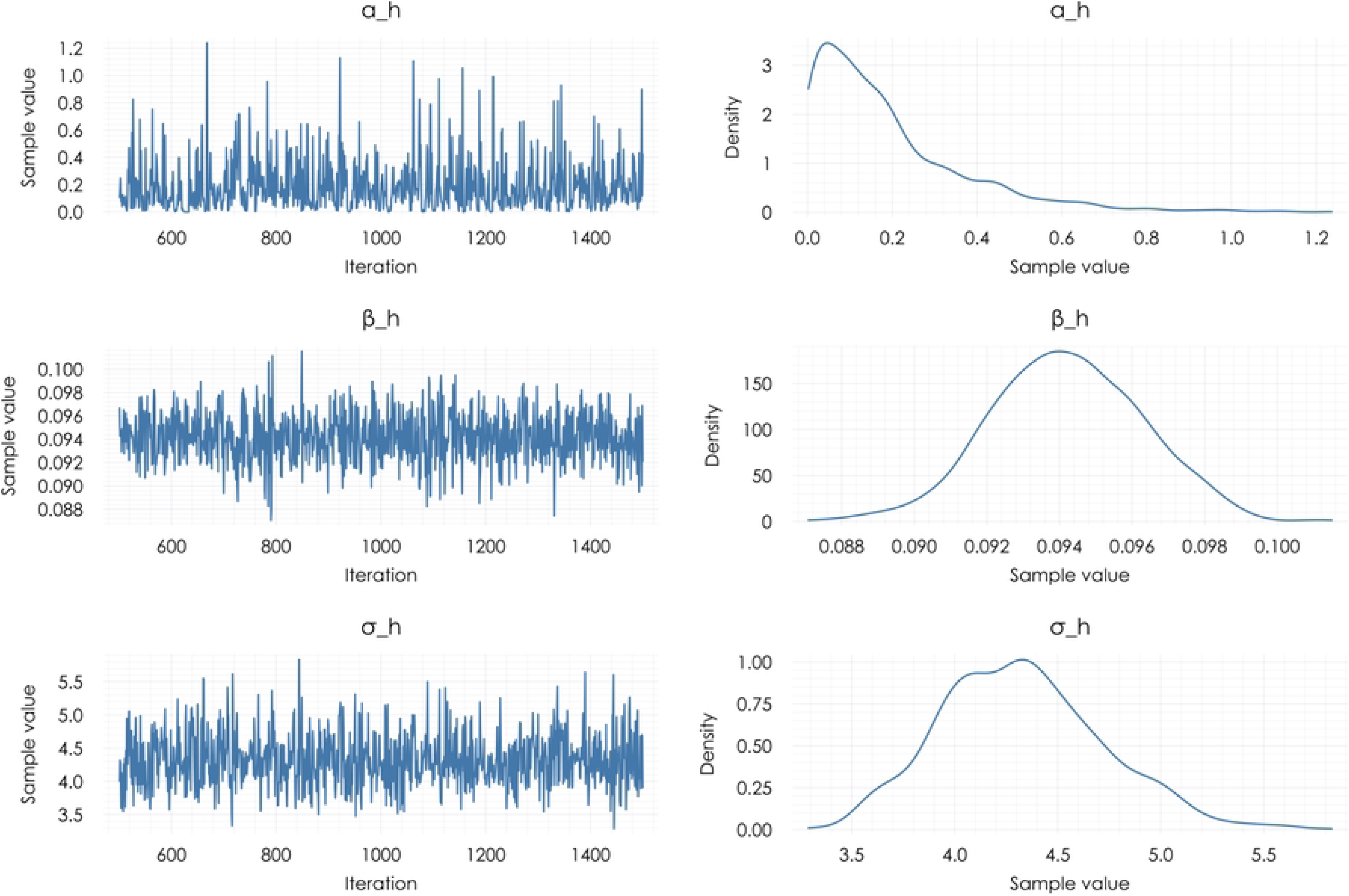
Posterior distributions of regression parameters. Probability Distribution of intercept (*α_h_*), slope (*β_h_*) and observation noise (*σ_h_*) after 1000 iterations (after 500 warm-up iterations) using NUTS

Using the *α_h_*, *β_h_* and *σ_h_*(and also the *α_u_*, *β_u_* and *σ_u_*) probability distributions, we can calculate a probabilistic line to visualise the accuracy of regression in the Thuringia dataset. Figure 7 shows these lines (top panel) as well as the predictions for hospitalisations and ICU cases (bottom panel).

**Figure 7.**
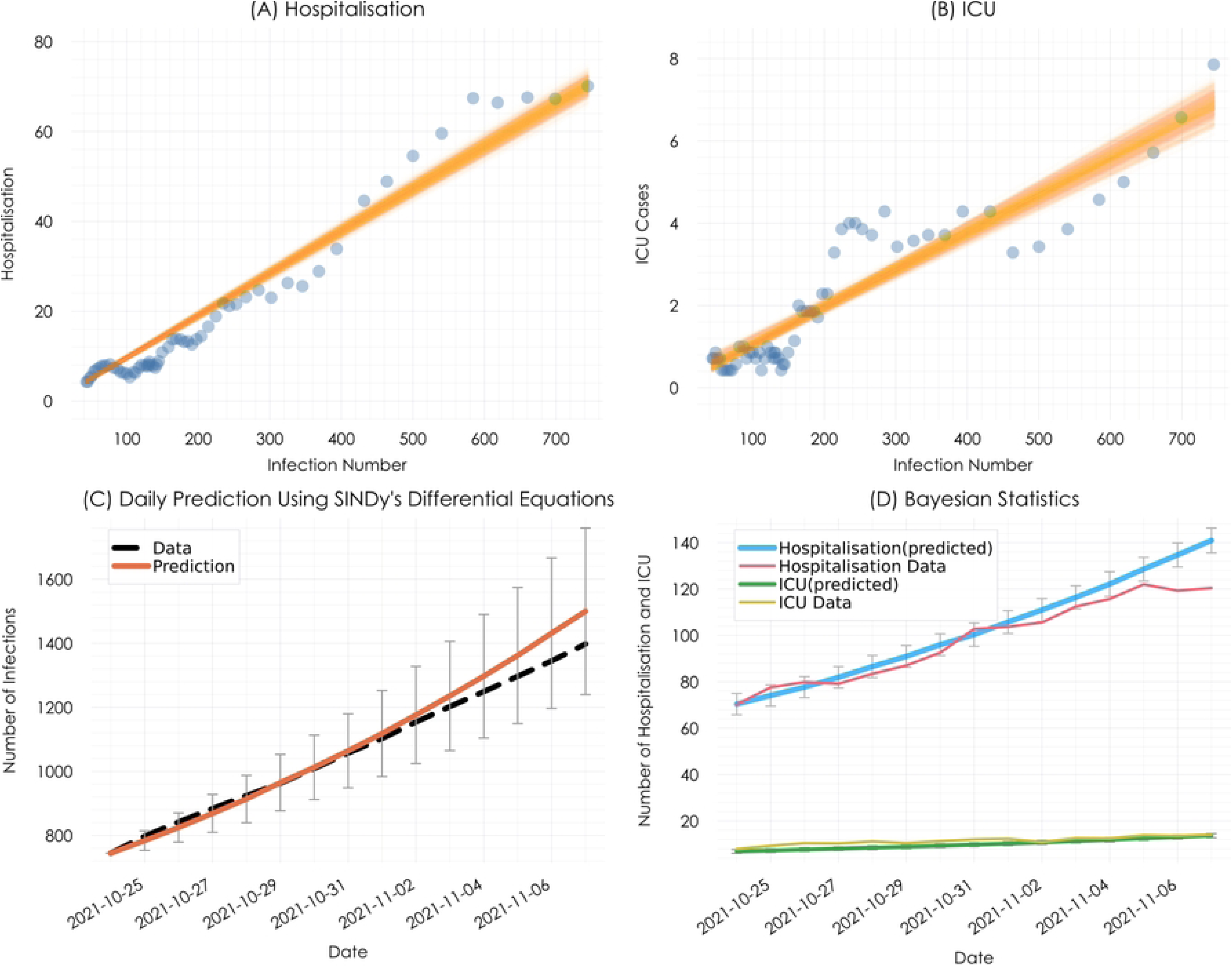
Using Bayesian statistics to predict hospitalisations and ICU cases. The panels on top (A and B) show the probabilistic lines. The orange line indicates the fit and +/− one sigma, and the panels on the bottom (C and D) show the predictions.

### A Useful Query

Since the control signal in the mathematical model is the antibody (obtained from vaccinations), we can feed the model with any arbitrary vaccination rate to see what happens in different scenarios. Some questions, like “What would have happened if we had not vaccinated people at this timeframe?” can be answered by performing such simulations with modified parameter sets. For example, in Figure 8, there are four different scenarios:

1. actual vaccinations: prediction with actual vaccinations, which is relatively compatible with the data.
2. no vaccinations: no one is getting vaccinated from 35 days before the prediction’s start day until the end of the prediction dates, which shows a sharp increase in the infections. According to the antibody kernel function [equation (4)], this scenario means zero antibodies during the prediction days.
3. start vaccinations: no further vaccinations from 35 days before the prediction date up to the start of the prediction dates, then just from the first day of prediction, vaccinations start at the actual rate (equal to actual vaccinations). The result of this scenario first shows an increase in the infections, but after a week, it starts to decrease because of the gradual accumulation of the antibody.
4. stop vaccinations: this scenario has the actual vaccination rate before the prediction day, but after that, no further vaccinations are given during the prediction dates. The result initially shows a similar result to the scenario one, but gradually, the infections increases due to the lack of vaccinations. This figure illustrates how our model uses the antibody (control signal) to incorporate the vaccinations history.

**Figure 8.**
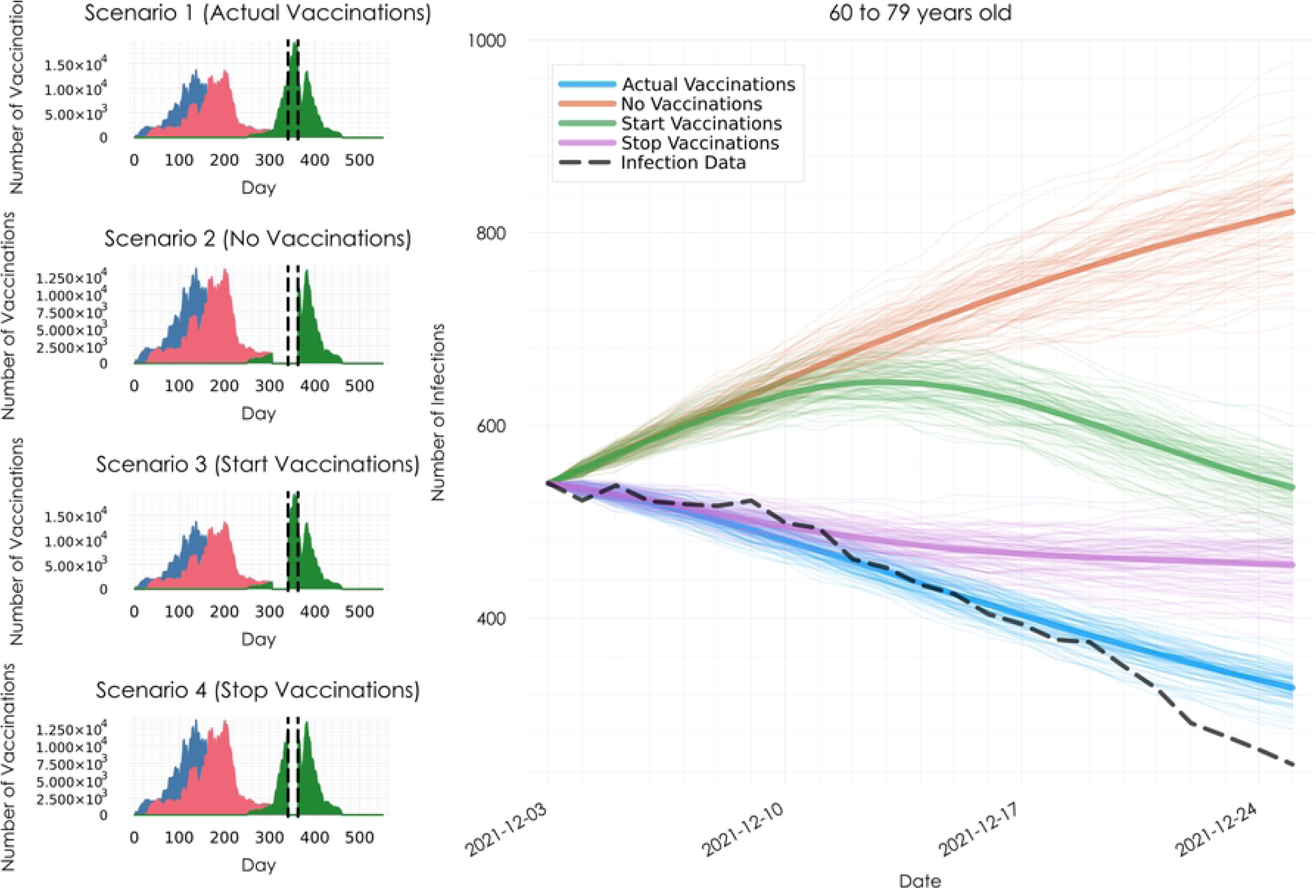
Predictions under different vaccination scenarios. The panels on the left show the vaccination inputs (blue: 1st dose, red: 2nd dose, light green: 3rd dose). The black vertical lines indicate the time window corresponding to the predictions shown in the right-hand panel. The right-hand panel displays the predicted number of infections for each scenario, compared with observed infection data.

The comparison of the results across different scenarios shows that the vaccination campaign had a significant effect in reducing infections, as any decrease in the vaccination rate leads to an increase in the number of positive cases.

### Coefficient Optimization

The governing differential equations of the pandemic are extracted from data using SINDy. These equations include several coefficients, which are influenced by external factors (like public health interventions, medical breakthroughs, changes in virus strain, etc.) over time. Optimising these coefficients helps us understand the effects of such factors on the pandemic and identify moments when significant changes occurred. We refer to the coefficients in the original ODE extracted by SINDy as global coefficients. As explained before, we use three approaches to optimise the global coefficients:

- Local coefficient adjustment
- Time-dependent coefficient adjustment
- Neural-augmented ODE adjustment

In this part, we want to show how practical each approach is. Table 3 shows the parameters used in optimisation for the two first approaches (local coefficient adjustment and time-dependent coefficient adjustment). All of the parameters are identical except the penalty function (see Equations 7 and 8) and the fact that in the local coefficient adjustment, we used seven days of data before the prediction day to adjust the coefficients and then made predictions, but in the time-dependent coefficient adjustment, we optimised the coefficients for every single day during the pandemic and then used the prediction start day’s coefficients to make predictions. In the local coefficient adjustment, the DiffEqParamEstim.jl package [26] was used to optimise the coefficients, but in the time-dependent coefficient adjustment, we used the Optim.jl package [27].

**Table 3.** Optimisation process parameters for local and time-dependent coefficient adjustment.

| Parameter | Value |
| --- | --- |
| **Loss Function** | Mean Squared Error (MSE) |
| **Differentiation** | Forward Differentiation |
| **ODE Solver** | Tsitouras 5<sup>th</sup> Order Runge-Kutta (Tsit5) |
| **Iterations** | 10<sup>3</sup> |
| **Optimiser** | Broyden-Fletcher-Goldfarb-Shanno (BFGS) |
| Penalty Function (Local Coefficient Adjustment) | Squared Euclidean Distance |
| Penalty Function (Time-dependent Coefficient Adjustment) | Total Variation Regularisation (TVR) |

To check the efficiency of the approaches, we considered a date on which the global coefficients perform poorly, i.e. producing high residuals. An ideal test case is a day in the middle of a sharp surge in infections, when authorities typically intervene with restrictions. Figure 9 shows the data and the results using three different sets of coefficients. The vertical line is the start day of the prediction, and the seven days before this line were used for the local coefficient adjustment. As it is evident from Figure 10, the global coefficients’ output fails to make a good prediction. After the local coefficient adjustment, the prediction is better, but still deviated from the data. The third approach is the time-dependent coefficient adjustment, in which we managed to capture almost all external factors, and the result matches the infections data.

**Figure 9.**
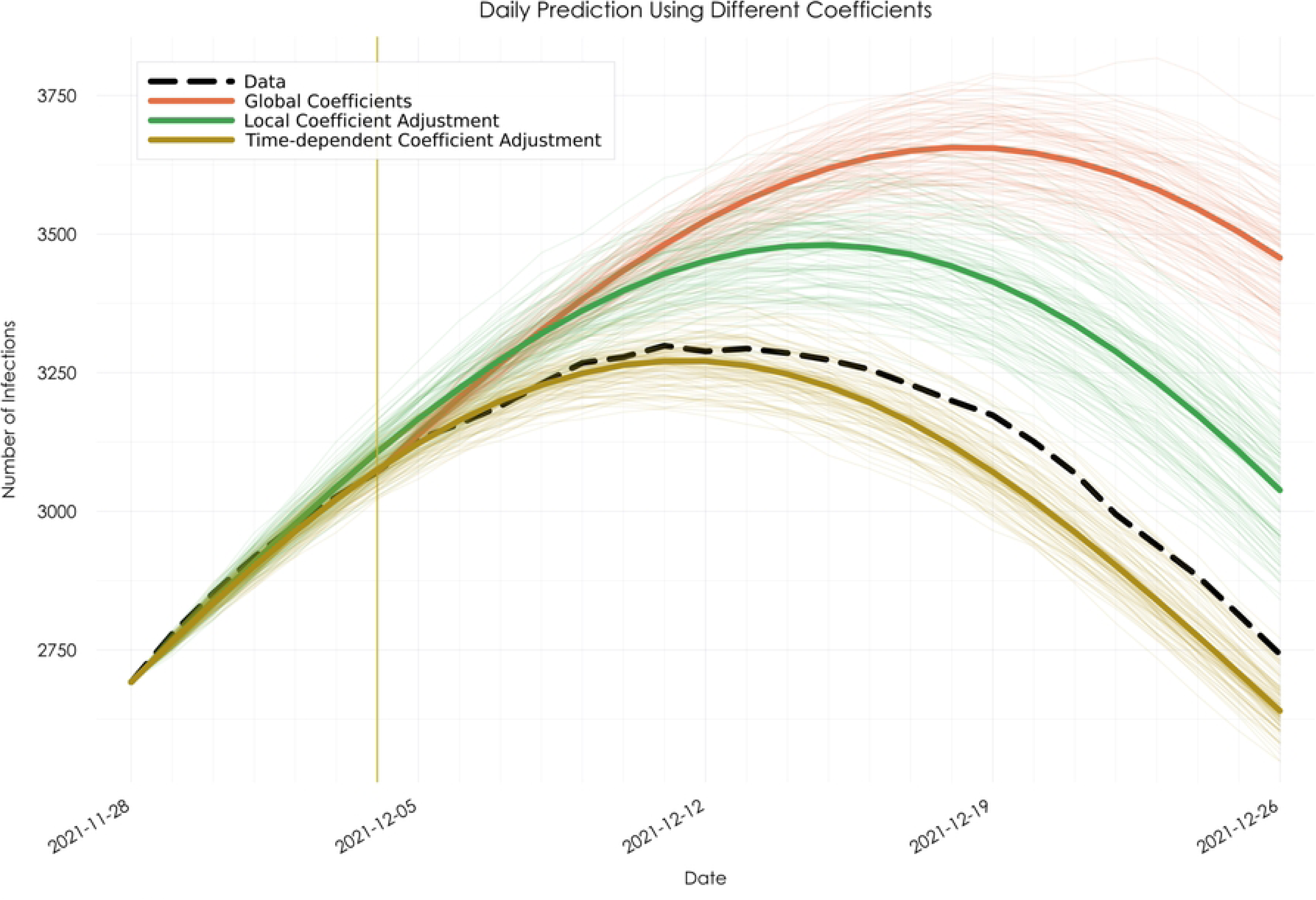
Daily infection predictions using three coefficient strategies. The dashed black line shows observed infections. The orange curve uses global coefficients, the green curve uses locally adjusted coefficients (based on the previous seven days), and the yellow curve applies time-dependent coefficients. The vertical red line marks the prediction start date.

**Figure 10.**
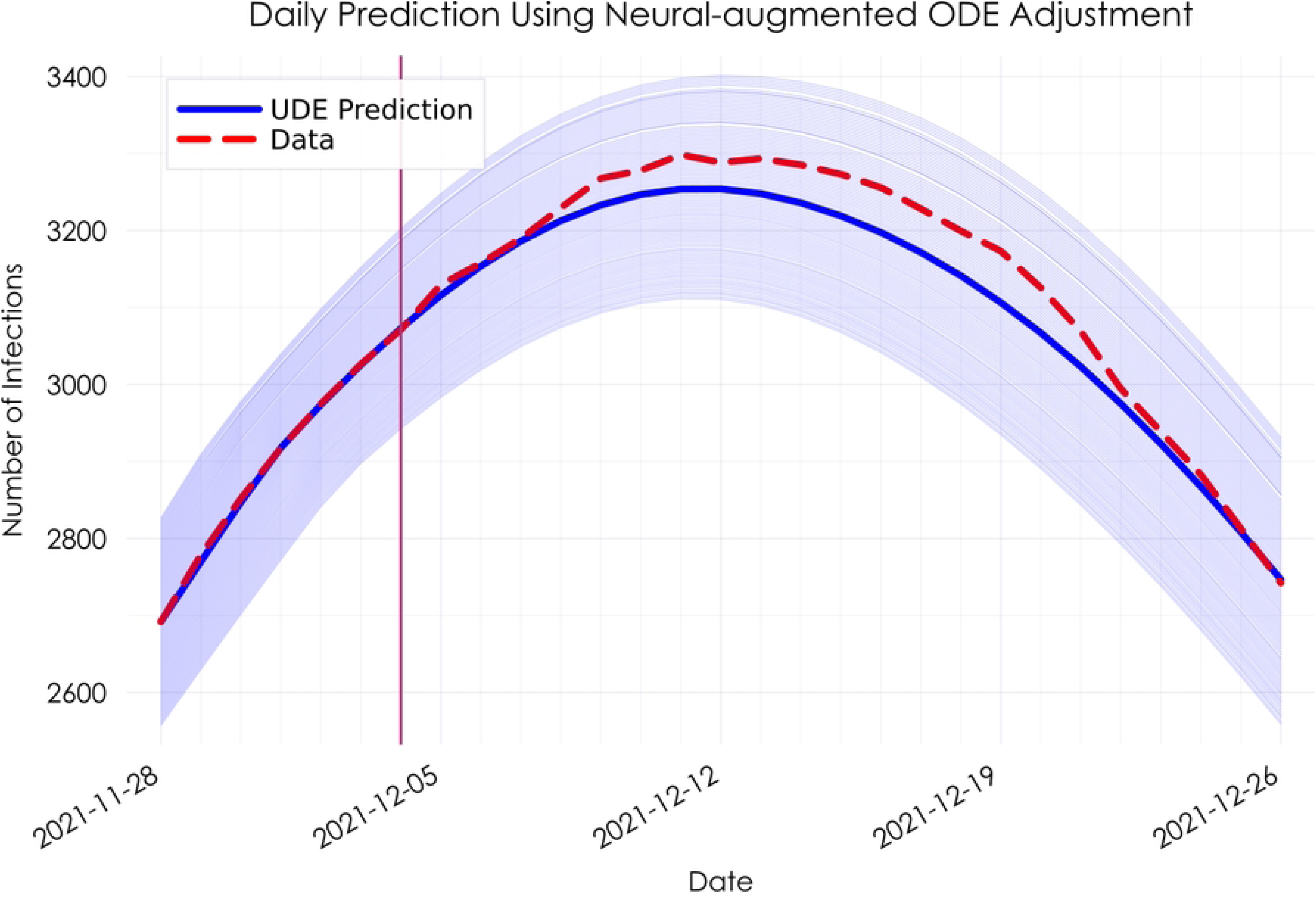
Infection prediction with neural-augmented ODE approach. The Universal Differential Equation (UDE) model, within the neural-augmented ODE adjustment approach, was used to forecast daily infections. The solid blue line is the prediction, and the shaded area shows uncertainty from ±5% variation in initial values. The red vertical line marks the prediction start date.

The third approach is the neural-augmented ODE adjustment, implemented using the universal differential equation (UDE) framework. Table 4 shows the parameters used in the optimisation process. This optimisation is carried out in two steps. The adaptive moment estimation (ADAM) is used in the first step with a learning rate 0.001. In the second step, the output of ADAM will be further optimised using Broyden-Fletcher-Goldfarb-Shanno (BFGS). The Lux.jl package[28] was used to create the neural network part and the Optimisation.jl [27] package was used to optimise the UDE.

**Table 4.** Optimisation parameters for the neural-augmented ODE adjustment, using UDE.

| Parameter | Value |
| --- | --- |
| Loss Function | Mean Squared Error (MSE) |
| **NN Structure** | 3x5x5x5x5x3 |
| **Activation Function** | The Radial Basis Function (RBF) |
| **Iteration** | $10^3$ |
| **ODE Solver** | Tsitouras 5th Order Runge-Kutta (Tsit5) |
| **Optimiser 1** | Adaptive Moment Estimation (ADAM) |
| **Learning Rate Optimiser 1** | 0.001 |
| **Optimiser 2** | Broyden-Fletcher-Goldfarb-Shanno (BFGS) |

Figure 10 shows the optimisation results for the same frame of time used in Figure 9 The vertical line is the start day of the prediction. The days before this line (seven days) were used to train the neural network in the UDE. The prediction is closely aligned with the observed data, indicating that external factors have been well captured. To test robustness, we simulated with ±5% variation in the initial infection values. The resulting predictions remain stable and consistent, as shown by the shaded blue region.

### Model Evaluation and Validation

After employing three different optimisation methods, we have four options for prediction, global coefficients, local coefficient adjustment, time-dependent coefficient adjustment, and neural-augmented ODE adjustment. To assess all of them, we utilised the walk-forward validation (WFV) method, which functions similarly to cross-validation in time-series data [29]. During each evaluation round, a data window is segmented as test data, initially positioned at the start and progressively moved forward through the dataset. Our window spanned 14 days, or two weeks, and shifted forward by one day in each step. Figure 11 displays the average percentage of the absolute residuals. The results indicate that the time-dependent coefficient adjustment captures temporal external factors most effectively, although the neural-augmented ODE adjustment performs slightly better towards the end of the second week. A key observation is that, across all optimisation methods, the residuals initially improve but then gradually increase, nearing the performance of the global coefficients. This trend suggests that these optimisation methods effectively capture temporal factors, making them well-suited for short-term predictions.

**Figure 11.**
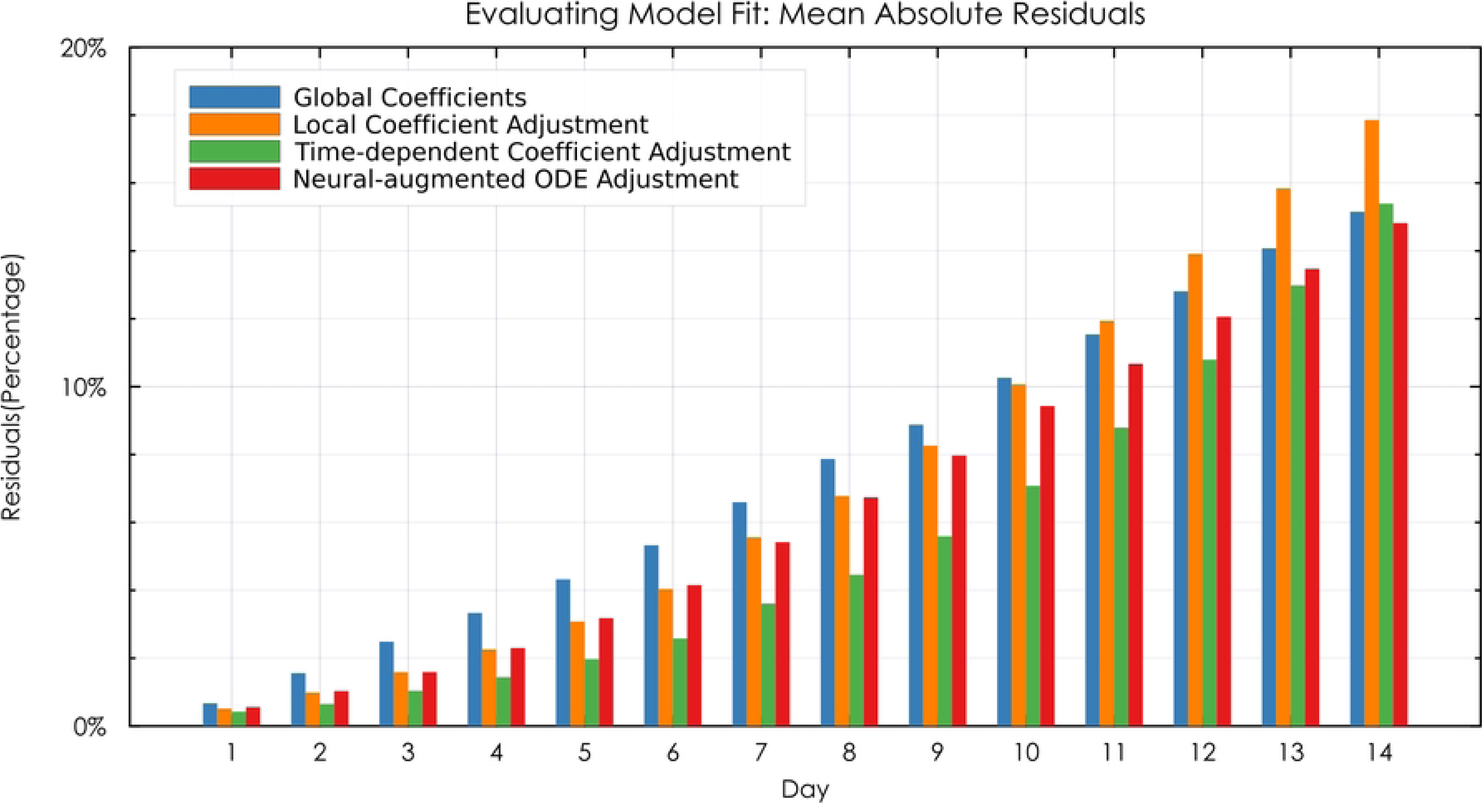
Mean absolute residuals (%) over a 14-day walk-forward validation (WFV) period. It is comparing four prediction strategies: global coefficients, local coefficient adjustment, time-dependent coefficient adjustment, and neural-augmented ODE adjustment. Time-dependent and neural-augmented models yield lower residuals, with the latter performing slightly better toward the end of the forecast window. Residuals increase over time for all methods, highlighting their strength in short-term prediction.

Table 5 presents three key metrics—*RMSE* (Root Mean Squared Error)*, MAE* (Mean Absolute Error)*, and R^2^* (coefficient of determination)—of the predictive models over time intervals of three, seven, ten, and fourteen days, calculated based on the number of infections. *RMSE* and *MAE* are standard measures of prediction error, with RMSE giving higher weight to larger errors due to squaring, while *MAE* provides a more uniform assessment of average error magnitude. *R²* indicates how well the model explains the variance in the observed data, with values closer to 1 representing better fit. The results indicate that the time-dependent coefficient adjustment approach achieves the highest accuracy, capturing over 90% of the predictive variance by the tenth day (*R^2^ = 0.91*) and maintaining strong performance at two weeks (*R^2^ = 0.83*). However, for predictions extending beyond two weeks, the neural-augmented ODE adjustment demonstrates superior outcomes, as shown in Figures 12 and 13, offering more reliable predictions in scenarios of higher infection rates. Given that daily infection rates in Thuringia can reach up to 4,000 cases, the observed values of *RMSE* and *MAE* provide a robust basis for reliable two-week forecasts

**Table 5.** Comparative performance metrics of predictive approaches over four different time intervals based on the infections.

| Model | Global coefficients |  |  | Local coefficient adjustment |  |  | Time-dependent coefficient adjustment |  |  | Neural-augmented ODE adjustment |  |  |
| --- | --- | --- | --- | --- | --- | --- | --- | --- | --- | --- | --- | --- |
| day | R<sup>2</sup> | RMSE | MAE | R<sup>2</sup> | RMSE | MAE | R<sup>2</sup> | RMSE | MAE | R<sup>2</sup> | RMSE | MAE |
| 3<sup>rd</sup> | 0.75 | 14.95 | 11.27 | 0.89 | 9.92 | 7.55 | 0.95 | 6.98 | 5.59 | 0.9 | 9.63 | 7.25 |
| 7<sup>th</sup> | 0.56 | 46.92 | 38.21 | 0.78 | 33.19 | 26.48 | 0.91 | 21.7 | 17.74 | 0.8 | 32.34 | 25.82 |
| 10<sup>th</sup> | 0.42 | 75.88 | 62.06 | 0.59 | 63.69 | 49.52 | 0.83 | 41.18 | 32.36 | 0.66 | 58.33 | 46.04 |
| 14<sup>th</sup> | 0.22 | 119.08 | 97.57 | 0.22 | 119.06 | 91.33 | 0.61 | 84.51 | 63.30 | 0.45 | 99.97 | 78.88 |

**Figure 12.**
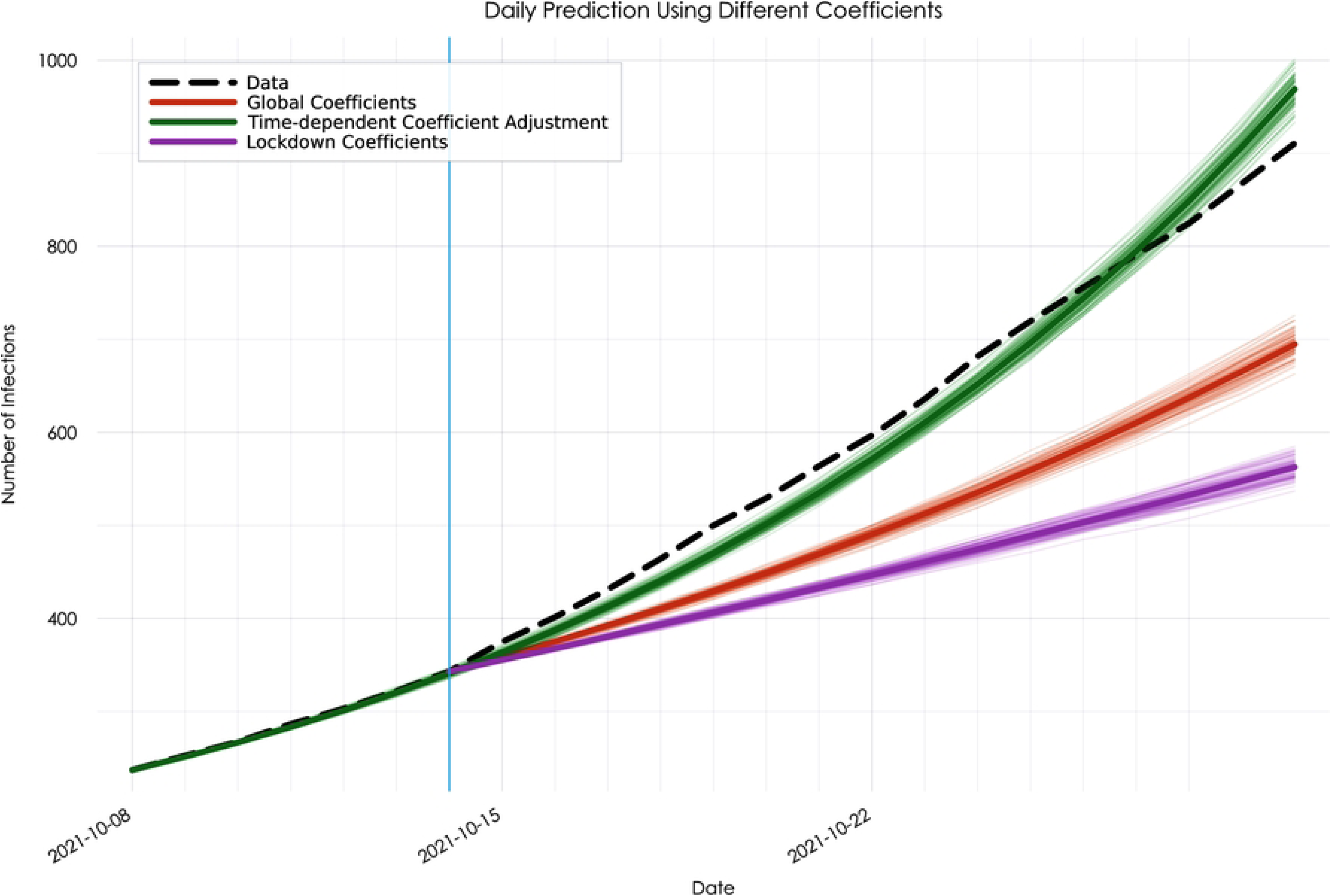
Daily infection predictions using different coefficient strategies. The dashed black line shows the infections data. The orange curve represents the prediction using global coefficients. The green curve shows the result of time-dependent coefficient adjustment, using the corresponding coefficients at the prediction date. The purple curve illustrates the scenario where coefficients associated with a past lockdown period are used instead of the actual time-dependent ones.

**Figure 13.**
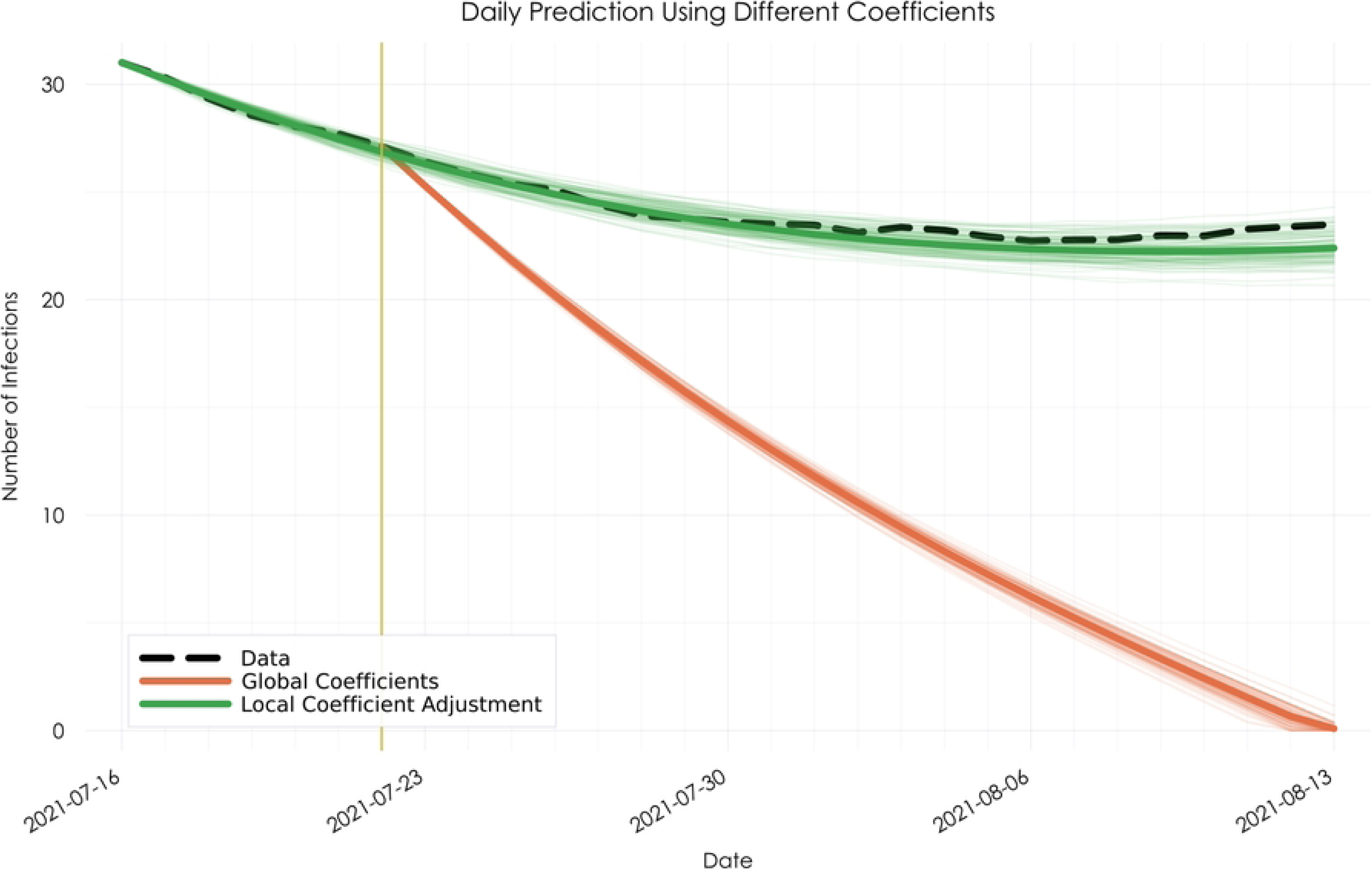
Daily infection predictions using global and locally optimised coefficients in a low-incidence scenario. The dashed black line represents the observed infection data, which remains below 50 cases per day. The orange curve shows the prediction based on global coefficients, which fails to capture the trend and drops unrealistically to zero. In contrast, the green curve, generated using the local coefficient adjustment approach, follows the observed trajectory closely. The vertical yellow line marks the start of the prediction period.

### Optimisation Insights

As outlined above, we employ three distinct strategies to optimise the global coefficients. This raises the question: why are multiple methods necessary, rather than relying on a single approach? The answer lies in the fact that each optimisation strategy offers its own advantages and is suited to different scenarios. While the neural-augmented ODE adjustment generally yields the most favourable outcomes for two-week prediction, the local coefficient adjustment and the time-dependent coefficient adjustment offer their own advantages.

Following the time-dependent coefficient adjustment, we obtain time-dependent coefficient values, represented as an array denoted as *c*, which encapsulates the optimal coefficients for each pandemic day. Leveraging these coefficients, we not only enhance prediction accuracy by utilising the coefficients specific to each day (*c_i_* for date *i*) but also facilitate scenario analysis. We can simulate the potential outcomes of interventions initiated from the respective dates by employing coefficients from specific dates, such as during a lockdown. This provides valuable insights into the effectiveness of interventions throughout different stages of the pandemic. Figure 12 demonstrates the application of the time-dependent coefficient adjustment. The vertical line marks the prediction starting day, with three predictions presented: the orange and green lines represent predictions made using the global coefficients and the time-dependent coefficient adjustment, respectively. It is evident from the figure that the time-dependent coefficient adjustment yields more accurate predictions than the global coefficients. The bottom line depicts the potential effects of an intervention. By utilising coefficients observed during previous lockdowns, it illustrates the potential outcome if a lockdown were to be initiated from the prediction starting day.

The local coefficient adjustment is another approach we used to optimise the coefficients of the global coefficients. While the time-dependent coefficient adjustment and the neural-augmented ODE adjustment yield superior results overall, our study reveals a notable advantage of the local coefficient adjustment in scenarios with extremely low infections. Throughout our analysis of the Thuringia pandemic dataset, where infection numbers typically ranged from 0 to 4000, we encountered challenges when daily cases dropped below 50. In such instances, global coefficients often faltered, yielding unreliable predictions. However, through the application of the local coefficient adjustment, we achieved efficient calibration of coefficient values, enabling the model to forecast future trends accurately despite minimal infection rates. Quantitative comparisons with the two other optimisation approaches underscored the efficacy of the local coefficient adjustment in these circumstances, demonstrating its value as a complementary tool for predictive modelling in the COVID-19 pandemic. Figure 13 shows the model’s outputs using the global coefficients and the local coefficient adjustment. The vertical line shows the prediction starting day. The seven days before this line were used in the optimisation process.

### What we learn from the model

By looking at this differential equation extracted by SINDy (Equation 11) we can gain a better insight into the dynamics of the pandemic. Here, *x* represents the daily number of infections, *y* represents the infectiveness (a convolution of *x*), and *A* shows the antibody effect from the vaccinations (convolution of the vaccinations *v*).

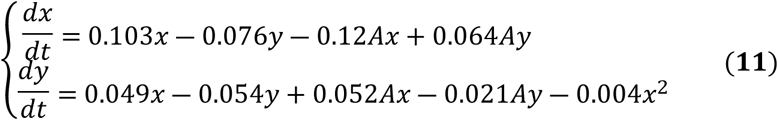

This model, derived directly from the data, highlights key dynamics of the pandemic. The first equation suggests that while infections (*x*) grow naturally at a base rate (0.103), they are suppressed by infectiveness (*-0.07y*) and further mitigated by vaccination (*-0.12Ax*). The relatively small suppressing effect of infectiveness might suggest a decrease in the susceptible population. Interestingly, the term *Ay* positively contributes to the infections, potentially reflecting partial vaccine efficacy. The second equation captures the dynamics of infectiveness (*y*). Since it is the convolution of the infections, it grows according to the infections (*0.049x*) and then decays over time (*-0.054y*). Vaccination in this equation exhibits a dual effect: On the one hand, it suppresses infectiveness (*-0.021Ay*) which is the direct biological effect of the vaccinations, on the other hand it amplifies the effect of the infections on the infectiveness (*0.052Ax*), which is an indirect impact of the vaccinations by making people more relaxed and increasing the transmission potential. The nonlinear term *-0.004x^2^* suggests a saturation effect at high infection levels, indicative of population-level herd immunity and it may also reflect underreporting when health institutions are overwhelmed. To deepen our understanding of the model’s dynamics, we now turn to its sensitivity analysis.

The sensitivity analysis of our model (Figure 14) shows that the infections coefficient, which scales with the number of active infections, has a strong effect on how the outbreak progresses. This impact is most noticeable at the beginning or middle of a new wave, where even a small reduction in the number of infectious individuals can lead to a significant drop in the future cases. This finding suggests that isolating infected individuals is one of the most effective ways to control the virus early on, as it quickly lowers the number of people spreading the infection. On the other hand, during the decline phase, changes in the infection rate have less effect, and at the peak of a wave, the model becomes unstable under sensitivity analysis, making predictions less reliable. Overall, this analysis highlights the importance of isolation as a key strategy to slow the spread, especially when cases are starting to rise.

**Figure 14.**
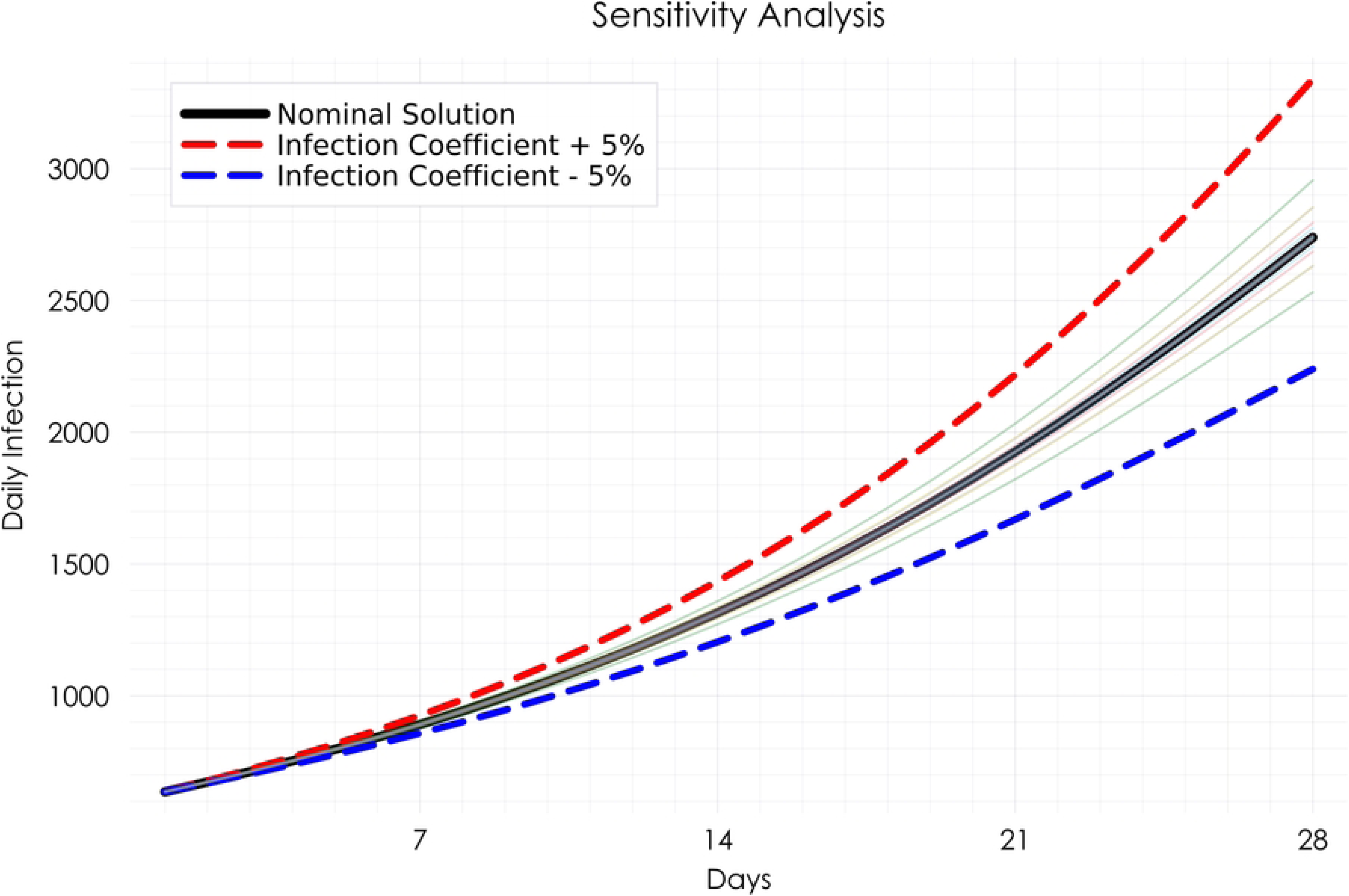
Sensitivity analysis highlighting the dominant influence of the infection coefficient in the ODE model. The black line shows the nominal prediction of daily infections. The red and blue dashed lines represent the model output when the infection coefficient is increased or decreased by 5%, respectively. The faded lines illustrate the effect of similar ±5% perturbations applied to other model coefficients. The visibly larger spread caused by changes in the infection coefficient indicates its critical role in driving the epidemic dynamics.

## Discussion and Conclusion

In conclusion, the results obtained from the comprehensive analysis of Thuringia’s COVID-19 dataset have provided valuable insights into the dynamics of the pandemic within the region. Through the utilisation of sophisticated mathematical models and optimisation techniques, we have been able to capture and predict various aspects of the pandemic, ranging from infection spread to hospitalisation rates. The impact of different vaccination scenarios’ effectiveness and external factors on infection dynamics has been thoroughly examined. Furthermore, the comparison of optimisation approaches has highlighted the strengths and limitations of each method, offering a nuanced understanding of their applicability in different scenarios. Overall, these findings contribute to a better understanding of COVID-19 dynamics and provide valuable tools for decision-making and intervention planning in managing the ongoing pandemic.

This research has extensively employed advanced mathematical modelling and optimization techniques to scrutinise the dynamics of the COVID-19 pandemic using data from Thuringia, Germany. The application of the Sparse Identification of Nonlinear Dynamical Systems (SINDy) algorithm, along with rigorous optimisation approaches, has enabled a nuanced understanding of infection propagation, vaccination effects, and public health interventions.

Key to our findings was the development of a robust model that integrates newly derived features (infectiveness and antibody) through sophisticated data processing techniques. This integration has significantly enhanced the accuracy of our predictions, providing deeper insights into the temporal dynamics of the virus’s transmission and the effectiveness of vaccination campaigns.

The exploration of various vaccination scenarios has underscored the critical role of targeted immunisation strategies in controlling the spread of the virus. These scenarios have demonstrated how different policy interventions can significantly alter infection trajectories, thus offering valuable lessons for public health strategy and response planning. Importantly, they also highlight that such interventions require time before their effects on infection dynamics become visible.

Moreover, our comparative analysis of different optimisation approaches, local coefficient adjustment, time-dependent coefficient adjustment, and neural-augmented ODE adjustment, highlights the distinct advantages of each method in capturing the complex interplay of epidemiological data and external factors influencing pandemic trends.

Despite their success in reflecting pandemic dynamics, all the models we applied are based on ODEs, which lack the hidden states used in methods like hidden Markov models or Kalman filters. While we incorporate historical context through the convolution of the infections and vaccinations, our approach primarily relies on data at individual time points and optimized coefficients from prior periods. This simplicity may limit predictive accuracy compared to models that explicitly handle latent states, such as exponential curve fitting or Kalman filters. However, by avoiding hidden states, our approach offers the advantages of easier interpretation and effective scenario analysis. Together, these models reveal the ongoing challenge of accurately modelling such complex systems. Future research should focus on refining these models to better account for emerging virus variants and shifts in public behaviour. Moreover, interdisciplinary approaches incorporating behavioural science and economics insights could enhance the model’s ability to simulate more realistic human responses to public health policies.

Our study not only advances the field of epidemiological modelling by providing a sophisticated tool for predicting disease spread but also offers a framework for policymakers to craft more effective and timely responses to public health crises. As we continue to face the challenges of COVID-19 and other infectious diseases, the insights gained from this research will be crucial in guiding future efforts to protect public health and safety.

## Acknowledgements

This work was supported by the German Federal Ministry of Research, Technology and Space within the funding program Photonics Research Germany (contract number 13N15742). We also thank the Scientific Advisory Council of the Free State of Thuringia for providing access to the dataset.

## Data Availability

The raw data underlying this study cannot be made publicly available because legal and data-protection regulations do not permit public access to the original reports and their data structures. However, the processed time-series data used in our analyses, as well as all code required to reproduce the results, are freely accessible in the project repository at: https://github.com/MortezaBabazadehShareh/Covid-19_modelling

